# Long-term temporal trends in incidence rate and case fatality of sepsis and COVID-19-related sepsis: nationwide registry study

**DOI:** 10.1101/2022.11.18.22282501

**Authors:** Nina Vibeche Skei, Tom Ivar Lund Nilsen, Siri Tandberg Knoop, Hallie C. Prescott, Stian Lydersen, Randi Marie Mohus, Alen Brkic, Kristin Vardheim Liyanarachi, Erik Solligård, Jan Kristian Damås, Lise Tuset Gustad

## Abstract

**Importance:** Sepsis is one of the leading causes of morbidity and mortality. The majority of sepsis cases is attributed to bacterial infections, but virus infections can also induce sepsis. Conflicting results in incidence rates and case fatality trends of sepsis is reported, and how the COVID-19 pandemic influenced these trends are unknown.

**Objective:** To estimate temporal trends in incidence rate and case fatality during a 14-year period from 2008 through 2021, and to assess possible shifts in these trends during the COVID-19 pandemic.

**Design:** A nationwide longitudinal registry study using ICD-10 discharge codes to identify sepsis.

**Setting:** All Norwegian hospitals from 2008 through 2021.

**Participants:** All sepsis cases included 317.705 patients and of these, 222.832 had a first sepsis episode.

**Main outcomes and measures:** Annual age-standardized incidence rates with 95% confidence intervals (CI). Poisson regression was used to estimate changes in incidence rates across time, and logistic regression was used to estimate odds ratios for in-hospital death.

**Results:** Among 12.619.803 adult hospitalizations, 317.705 (2.5%) patients met the sepsis criteria and 222.832 (70.0%) had a first sepsis episode. In the period 2009-2019, the annual incidence rate for a first sepsis episode was stable (incidence rate ratio per year, 0.999; 95% CI, 0.994-1.004), whereas for all sepsis the incidence rate increased by 15.5% during the period (annual incidence rate ratio, 1.013; 95% CI 1.007-1.019). During the COVID-19 pandemic, the incidence rate ratio for a first sepsis was 0.877 (95% CI, 0.829-0.927) in 2020 and 0.929 (95% CI, 0.870-0.992) in 2021, and for all sepsis it was 0.870 (95% CI, 0.810-0.935) in 2020 and 0.908 (95% CI, 0.840-0.980) in 2021, compared to the previous 11-year period. In-hospital deaths declined in the period 2009-2019 (odds ratio per year, 0.954 [95% CI,0.950-0.958]), whereas deaths increased during the COVID-19 pandemic in 2020 (odds ratios, 1.061 [95% CI 1.001-1.124] and in 2021 odds ratio (1.164 [95% CI, 1.098-1.233]).

**Conclusion and relevance:** We found a stable incidence rate of a first sepsis episode during the years 2009-2019. However, the increasing burden of all sepsis admissions indicates that sepsis awareness with updated guidelines and education must continue.

**Key Points:** *Question:* Has there been a change in incidence rate and case fatality of sepsis over the past decade, and how did the COVID-19 pandemic influence sepsis incidence rates and in-hospital mortality?

*Findings:* In this nationwide longitudinal registry study the incidence rate of all sepsis episodes increased and the incidence rate of a first sepsis episode was stable during the period 2009-2019, whereas in 2020 and 2021, the incidence rate of a first and all sepsis episodes was lower than in the preceding 11-year period. Case fatality risk declined from 2009 to 2019, but increased somewhat in 2020 and 2021, when 9.7% of first sepsis cases were identified as COVID-19 related sepsis.

*Meaning:* Despite a stable incidence rate of first-time sepsis admissions over time, the burden of sepsis is rising due to an increased rate of patients admitted multiple times with sepsis. The COVID-19 pandemic have had an impact on sepsis incidence rate and hospital mortality and needs further evaluation.

## Introduction

Sepsis is a dysfunctional immune response to infection that leads to acute life-threatening tissue damage and organ dysfunction.^1^ With an estimated 50 million cases and 11 million sepsis-related deaths in 2017, sepsis remains a major cause of worldwide morbidity and mortality.^2^ While sepsis may result from any infection, the majority of adult sepsis cases before the pandemic were attributed to bacterial infections, and viral sepsis was rare.^3-5^ During the COVID-19 pandemic, an unprecedented number of patients developed viral sepsis,^6-9^ with a high risk of co-infections and secondary infections that can aggravate the outcome.^10,11^ It is likely that public health efforts to reduce the spread of SARS-CoV-2, such as lockdowns, may also have influenced the spread of other communicable diseases contributing to the risk of sepsis.^12,13^ However, only one study has evaluated the impact of the pandemic on sepsis incidence rate and case fatality risk, using a few selected sepsis codes.^14^ No previous study has focused exclusively on sepsis incidence rate using all sepsis codes, ^2^ and compared sepsis incidence rate and case fatality during the two first years of the COVID-19 pandemic with long-term historic trends.

Previous research on the incidence rates of sepsis before the COVID-19 pandemic has shown conflicting results. ^2,15-17^ However, the incidence rate and mortality rate are challenged, and more accurate quantification (i.e., correct identification and diagnosis coding) of sepsis are warranted.^18,19^

The overall aim of this study is therefore to describe temporal trends in sepsis incidence rate and case fatality using nationwide data on all adult hospital admissions from 2008 through 2021, and secondly to examine changes in hospital admission and mortality rates of sepsis during the first two COVID-19 pandemic years.

## Methods

### Data Source and Study Population

This nationwide longitudinal study used data from the Norwegian Patient Registry (NPR) and Statistics Norway.^20,21^ NPR is an administrative database maintained by the Norwegian Directorate of Health that contains data with unique patient identifiers that allow follow-up of individual patients on every admission to public hospitals in Norway from 2008 onward. In addition, NPR contains admission and discharge dates, and the International Classification of Diseases 10^th^ revision (ICD-10) discharge codes, while Statistics Norway contains demographic data on all citizens of Norway. In NPR, we identified all hospitalizations to public hospitals in Norway (2008–2021) aged ≥18 years with the ICD-10 discharge diagnosis code(s) for sepsis consistent with the Angus implementation refined by Rudd and colleagues.^2,22^

We treated each hospitalization as an individual entry, and within this entry, sepsis was defined as explicit or implicit sepsis. For explicit sepsis, we used the presence of one code (See Supplementary Table 1 for an overview of all ICD-10 codes to define sepsis). For implicit sepsis, we used the combination of one infection code with the presence of an acute organ dysfunction code. The strategy was used for the primary and up to 20 secondary co-existing ICD-10 discharge codes since there is no obligatory order for the secondary codes. We added COVID-19-related sepsis to the implicit sepsis category based on the presence of a diagnostic code for COVID-19 (U07.1, U07.2) and ≥one organ dysfunction code. Patients with a COVID-19 sepsis code and an explicit sepsis code were categorized as explicit sepsis. Figure 1 shows the selection of patients into the study.

**Fig. 1.**
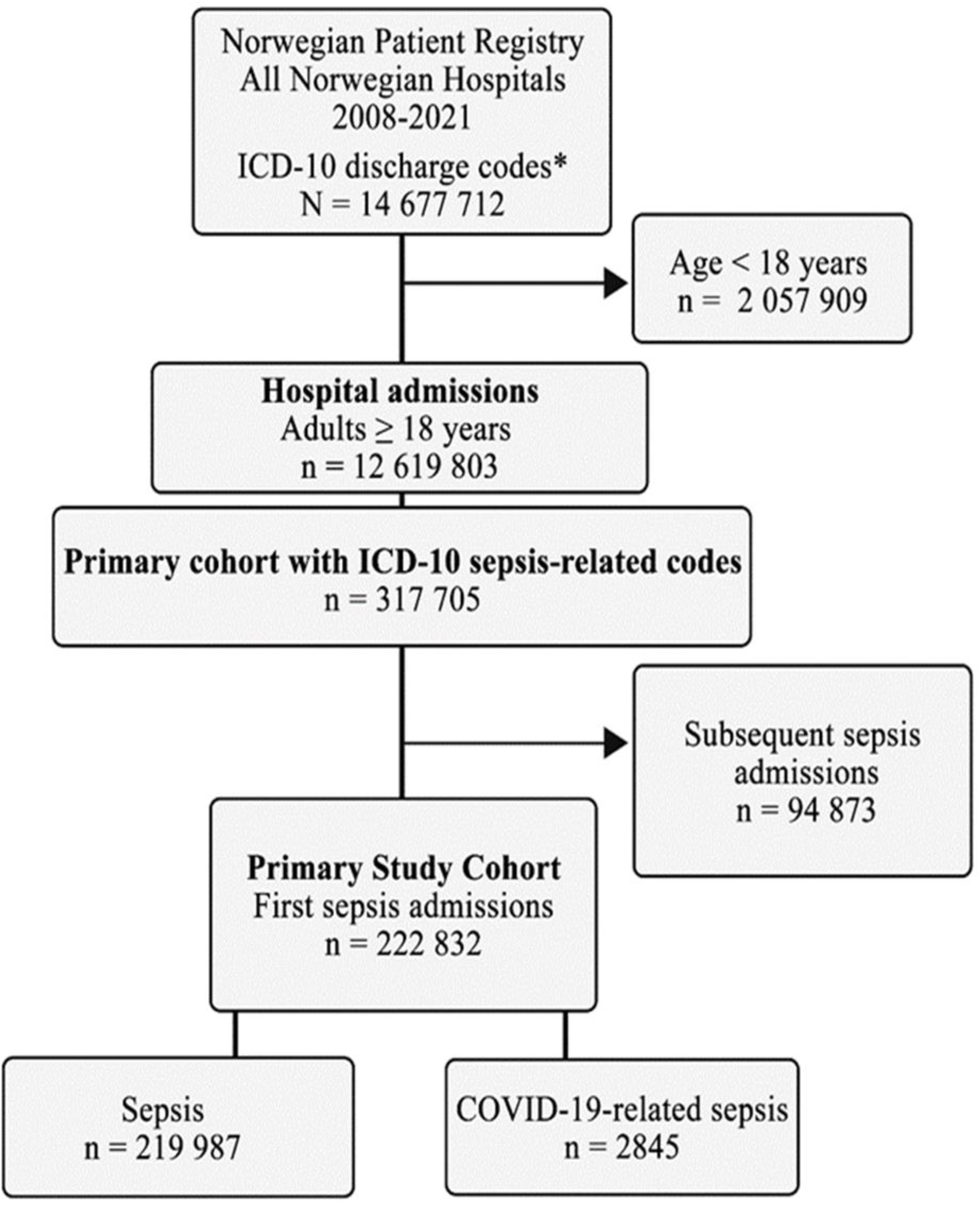
Flowchart of the study population with the inclusion and exclusion process. * = Non-psychiatric ICD-10 codes

### Characteristics of Study Population

Patient characteristics were extracted from NPR, including sex, age, ICD codes for selected comorbidities ^23^, as well as numbers of hospital stays from sepsis, readmissions, and in-hospital deaths (for details, see Supplementary Table 2). For sepsis admissions, we used ICD-10 codes to classify site(s) of infection into respiratory, genitourinary, intra-abdominal, extra-abdominal, endocarditis/myocarditis, soft tissue, infections following a procedure, and other (bone, joint, obstetric, ear, mouth, upper airway, central nervous system and unknown). The acute organ dysfunctions were classified by number and as circulatory, respiratory, renal, hepatic, coagulation, and/or other (acidosis, unspecific gangrene, and central nervous system.). Following Norwegian coding rules, hospitalizations with an infection code in combination with diagnostic code R65.1, Systemic Inflammatory Response Syndrome (SIRS), were not considered to have implicit sepsis, and R65.1 did then not count as one organ failure. However, R65.1 together with any other co-existing acute organ dysfunction code counted as one organ failure. A sepsis admission was defined as recurring sepsis admission if the patient was discharged with an explicit or implicit sepsis code and thereafter admitted with an explicit or implicit sepsis code, regardless of the time frame for the new admission. The number of sepsis admissions was categorized from one to five or more.

### Statistical Analysis

Descriptive statistics are presented as frequencies, means, standard deviation, percent, and medians as appropriate, and are reported by sepsis or COVID-19 status. We calculated the crude sepsis incidence rate (IR) of a first and overall sepsis episode according to year (2008–2021) and ten-year age-groups as the number of sepsis admissions divided by the total number of inhabitants in Norway at the beginning of the year. The IRs for first and all sepsis were then standardized according to Segi’s world standard population using ten-years age categories^24,25^, and reported per 100 000 person years.

To evaluate the temporal trends of sepsis incidence rates and the impact of the COVID-19 pandemic on sepsis incidence rates we used Poisson regression to estimate incidence rate ratios (IRR) of sepsis using the number of sepsis admissions (total or first) as the dependent variable, population as exposure, the years 2009 to 2019 as a continuous variable, and the years 2008, 2020 and 2021 as separate indicator variables. The analyses were adjusted for sex (man, woman) and age (10-year categories). Since 2008 was the first observation year, we could not differentiate between a first and a recurrent episode, and 2008 thus was included as an indicator variable to account for a possibly inflated incidence rate of first sepsis. To account for overdispersion, we used the robust variance estimator.

Case fatality risk (CFR) was estimated based on first sepsis admissions with a discharge status of in-hospital death divided by all first sepsis hospitalizations. The calculation was performed on annual cases from 2008 to 2021 and by ten-years age groups in the same period. During 2020 and 2021 we also calculated the quarterly CFR and compared CFR for COVID-19-related sepsis and sepsis. To evaluate the trend of in-hospital mortality and the pandemic’s impact on hospital mortality, we used logistic regression to estimate odds ratios (ORs) for in-hospital death using the years 2009-2019 as a continuous variable, the years 2008, 2020, and 2021 as indicator variables, and adjusting for sex (man, woman) and age (10-year categories). We report 95% confidence intervals (CI) where relevant.

All analyses were conducted using STATA version 16.1 (Stata Corp).

### Ethics

The study was approved by the Regional Committee for Medical and Health Research Ethics (REK) in Eastern Norway (2019/42772) and the Data Access Committee in Nord-Trøndelag Hospital Trust (2021/184). In accordance with the approval from the REK and the Norwegian law on medical research, the project did not require a written patient consent. This work was performed on TSD (Service for Sensitive Data) facilities owned by the University of Oslo, operated and developed by the TSD service group at the University of Oslo, IT Department (USIT). TSD is designed to store and post-process sensitive data in compliance with the Norwegian “Personal Data Act” and “Health Research Act.”

## Results

### Characteristics of Study Population

Among 12.619.803 non-psychiatric adult hospitalizations during the study period (2008–2021), 317.705 (2.5%) met the criteria for sepsis, and of these, 222.832 (70%) were hospitalized with the first episode of sepsis. Patient characteristics according to a first episode of sepsis and COVID-19-related sepsis are presented in Table 1.

**Table 1.** Characteristic of the study population at first sepsis admission (2008-2021) and COVID-19-related sepsis (2020-2021)

| Characteristics | Sepsis | COVID-19-related sepsis | All first sepsis admissions |
| --- | --- | --- | --- |
|  | n (%) | n (%) | n (%) |
| First admission n (% of all sepsis admissions) | 219 987 (69) | 2845 (1) | 222 832 (70) |
| **Sex** |  |  |  |
| Male | 118 580 (53.9) | 1862 (65.5) | 120 442 (54.1) |
| **Age (years)** |  |  |  |
| Mean ± SD (Median) | 71.2 ± 16.6 (74.4) | 61.4±16.1 (61.8) | 71.1 ± 16.6 (74.3) |
| **Number of comorbidities** |  |  |  |
| 0 | 66 869(30.4) | 1 581(55.6) | 68 450 (31.7) |
| 1 | 97 894 (44.5) | 909 (32.0) | 98 803 (44 .3) |
| 2 | 45 052 (20.5) | 300 (10.5) | 45 352 (20.4) |
| $\geq 3$ | 10 172 (4.6) | 55 (1.9) | 10 227 (4.6) |
| **Comorbidities <sup>a</sup>** |  |  |  |
| Heart and vascular | 99 360 (64.9) | 702 (55.5) | 100 062 (64.8) |
| Cancer | 39 243 (25.6) | 125(9.9) | 39 368 (25.5) |
| Lung | 35 859 (23.4) | 306 (24.2) | 236 165 (23.4) |
| Renal | 8 873 (5.8) | 76 (6.0) | 8 949 (5.8) |
| Diabetes | 24 030 (15.7) | 386 (30.5) | 24 416 (15.8) |
| Dementia | 8 068 (5.3) | 32 (2.5) | 8 100 (5.3) |
| Immune | 3 091 (2.0) | 49 (3.9) | 3 140 (2.0) |
| Liver | 991 (0.7) | NA | 994 (0.6) |
| **Site of infection <sup>b</sup>** |  |  |  |
| Respiratory | 79 290 (48.7) | 2 528 (97.9) | 81 818 (49.5) |
| Genitourinary | 44 700 (27.5) | 82 (3.2) | 44 782 (27.1) |
| Skin and soft tissue | 8 260 (5.1) | 5 (0.2) | 8 265 (5.0) |
| Intra-abdominal | 8 841(5.4) | 29 (1.1) | 8 870 (5.4) |
| Extra-abdominal | 12 318 (7.6) | 22 (0.9) | 12 340 (7.5) |
| Infections following a procedure | 8 277 (5.1) | 13 (0.5) | 8 290 (5.0) |
| Endocarditis/Myocarditis | 2 522 (1.6) | 8 (0.3) | 2 530 (1.5) |
| Other <sup>c</sup> | 28 836 (17.7) | 152 (5.9) | 28 997 (17.5) |
| **Explicit sepsis** | 77 240 (35.1) | 90 (3.2) | 77 330 (34.7) |
| **Number of Acute organ dysfunctions** |  |  |  |
| 1 | 131 310 (87.6) | 2 470 (89.1) | 133 780 (87.6) |
| 2 | 15 028 (10.0) | 270 (9.7) | 15 298 (10.0) |
| 3 | 2 851 (1.9) | 227 (1.0) | 2 878 (1.9) |
| $\geq 4$ | 695 (0.5) | NA | 700 (0.5) |
| **Organ system with acute organ dysfunction <sup>d</sup>** |  |  |  |
| Respiratory | 59 465 (41.0) | 2 399 (87.5) | 61 864 (41.8) |
| Circulatory | 14 824 (10.2) | 68 (2.5) | 14 892 (10.1) |
| Renal | 66 809 (46.1) | 433 (15.8) | 67 242 (45.5) |
| Hepatic | 3 192 (2.2) | 17 (0.6) | 3 209 (2.2) |
| Coagulation | 1 639 (1.1) | 12 (0.4) | 1 651(1.1) |
| Other<sup>e</sup> | 22 116 (15.2) | 151 (5.5) | 22 267 (15.1) |
| **Number of hospital admissions for sepsis <sup>f</sup>** |  |  |  |
| 1 | 168 904 (76.8) | 2 714 (95.4) | 171 618 (77.0) |
| 2 | 33 097 (15.0) | 4125 (4.4) | 33 222 (14.9) |
| 3 | 10 125 (4.6) | NA | 10 129 (4.6) |
| 4 | 40 010 (1.8) | NA | 4 011 (1.8) |
| $\geq 5$ | 3 851 (1.8) | NA | 3 852 (1.7) |
| **Readmission <sup>g</sup>** | 54 967 (25.0) | 474 (16.7) | 55 441 (24.9) |

If not mentioned otherwise, the percentage (%) is calculated from available data from the first admission with Sepsis or COVID-19-related sepsis.
Abbreviation: NA=Not Applicable (used when the number of admissions was $\leq 5$ ).
<sup>a</sup> The proportion of all comorbidities is calculated as number of particular comorbidity over total number of comorbidities
<sup>b</sup> The proportion of all infections sites is calculated as number of individuals with particular infection site over total number of infections sites
<sup>c</sup> Other infection sites= Bone, obstetric, upper airway, central nervous system and unknown
<sup>d</sup> The proportion of organ dysfunctions is calculated based on n with any organ dysfunctions
<sup>e</sup> Other acute organ dysfunction= Acidosis, unspecific gangrene and central nervous system dysfunctions.
<sup>f</sup> Number of hospital admissions= Calculated as new sepsis admission if admission with ICD-10 codes defining sepsis, regardless of time frame for the new sepsis admission. Follow up=14 years
<sup>g</sup> Readmission= admission within 30 days after discharge regardless of cause

In 2020 and 2021, 2.845 of 29.329 (9.7%) of first sepsis cases were identified as COVID-19 related sepsis. Men were overrepresented among patients with sepsis (53.9%) and COVID-19-related sepsis (65.5%). The sepsis patients were older than patients with COVID-19-related sepsis (mean age 71.1 vs. 61.4). The sepsis patients experienced renal acute organ dysfunction most often (46.1%). followed by respiratory failure (41.0%). The COVID-19-related sepsis patients experienced naturally most frequent respiratory failure (87.5%), followed by renal failure (15.8%). In total, 25.0% and 16.7% of the patients were readmitted within 30 days in the sepsis and COVID-19-related sepsis group, respectively. During the total study period (2008-2021), 24.2% of sepsis patients had ≥2 recurring sepsis hospitalization.

### Sepsis Incidence Rates and Temporal Trends

The overall age-standardized IR of a first sepsis admission was 246/100.000 (95% CI, 245-247), whereas the age-standardized IR of all sepsis admissions was 352/100.000 (95% CI, 351-354) during the study period (Table 2). The age-standardized IRs for first and all sepsis admissions by year 2008-2021 is given in Table 3 and Figure 2. More detailed information on IRs in different age groups is given in Supplemental Figure 1, showing the highest occurrence in the older age groups.

**Table 2.** Standardized incidence rates for first and all sepsis admissions 2008-2021^a^

| Year | No. of persons | Incidence rate first sepsis admission per 100 000 person years |  | Incidence rate all sepsis admissions per 100 000 person years |  |
| --- | --- | --- | --- | --- | --- |
|  |  | Crude | Adjusted (95% CI) | Crude | Adjusted (95% CI) |
| 2008 | 3 637 892 | 445 | 286 (281-291) | 526 | 344 (338-350) |
| 2009 | 3 697 780 | 401 | 257 (253-262) | 544 | 342 (336-347) |
| 2010 | 3 749 043 | 407 | 261 (257-266) | 546 | 357 (351-362) |
| 2011 | 3 805 931 | 402 | 260 (256-265) | 545 | 356 (351-361) |
| 2012 | 3 867 645 | 395 | 252 (247-256) | 553 | 358 (353-364) |
| 2013 | 3 928 378 | 380 | 240 (236-244) | 533 | 343 (337-348) |
| 2014 | 3 983 895 | 386 | 243 (238-247) | 555 | 352 (346-357) |
| 2015 | 4 040 198 | 401 | 250 (246-254) | 576 | 361 (355-366) |
| 2016 | 4 086 583 | 385 | 237 (233-241) | 577 | 359 (353-364) |
| 2017 | 4 127 266 | 409 | 246 (242-250) | 599 | 361 (356-366) |
| 2018 | 4 166 612 | 417 | 246 (242-250) | 622 | 367 (362-372) |
| 2019 | 4 205 704 | 409 | 240 (236-244) | 631 | 368 (363-373) |
| 2020 | 4 248 972 | 364 | 210 (206-213) | 561 | 322 (317-326) |
| 2021 | 4 279 679 | 390 | 226 (222-230) | 602 | 343 (338-348) |
| Total | 55 825 578 | 399 | 246 (245-247) | 569 | 352 (351-354) |

Abbreviation: CI = confidence interval
<sup>a</sup> Crude and age adjusted sepsis incidence rate was calculated by year (2008–2021) for first and all sepsis admissions by dividing sepsis admissions by the total number of inhabitants in Norway at beginning of the same years, using direct standardization weighted by 'Segi's world standard population.

**Table 3.** Poisson regression^a^ for trends of first and all sepsis episode

|  | First sepsis admissions |  | All sepsis admissions |  |
| --- | --- | --- | --- | --- |
|  | IRR | 95% CI | IRR | 95% CI |
| Per year 2009 to 2019 | 0.999 | 0.994-1.004 | 1.013 | 1.007-1.019 |
| 2008 | 1.110 | 1.021-1.210 | 1.007 | 0.920-1.102 |
| 2020 | 0.877 | 0.829-0.927 | 0.870 | 0.810-0.935 |
| 2021 | 0.929 | 0.870-0.992 | 0.908 | 0.840-0.980 |
| Female sex | 0.688 | 0.669-0.707 | 0.677 | 0.656-0.699 |
| Age group, years |  |  |  |  |
| 18-29 | 0.023 | 0.021-0.026 | 0.023 | 0.020-0.025 |
| 30-39 | 0.029 | 0.026-0.031 | 0.028 | 0.025-0.030 |
| 40-49 | 0.043 | 0.041-0.046 | 0.044 | 0.041-0.047 |
| 50-59 | 0.089 | 0.085-0.093 | 0.094 | 0.088-0.100 |
| 60-69 | 0.207 | 0.200-0.214 | 0.225 | 0.215-0.235 |
| 70-79 | 0.457 | 0.441-0.473 | 0.491 | 0.470-0.512 |
| ≥80 | 1.000 | Reference | 1.000 | Reference |
| Constant<sup>b</sup> | 0.031 | 0.030-0.033 | 0.040 | 0.038-0.042 |

Abbreviation: IRR = incidence rate ratio, CI = confidence interval
<sup>a</sup> The Poisson regression model was set up with cases as dependent variable, population as exposure, per year 2009-2019 as continuous covariate, and indicator variables as covariates for the years 2008, 2020 and 2021, and female sex and age groups.
<sup>b</sup> Constant = estimated incidence rate for men ≥80 in 2009

**Fig. 2.**
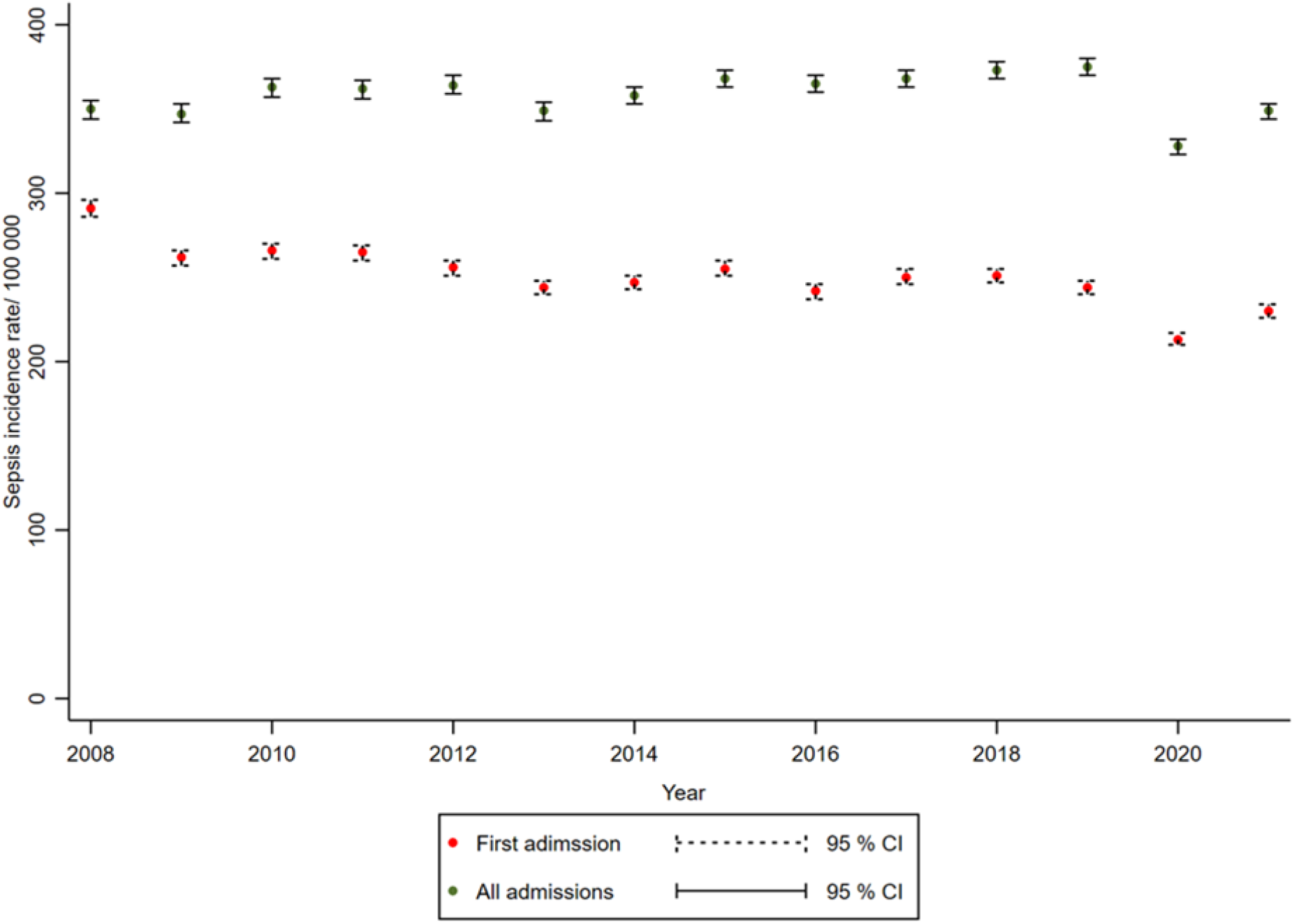
Annual standardized incidence rates of first and all sepsis admissions per 100 000 inhabitants. Abbrevation: CI = confidence interval

Poisson regression showed that from 2009 throughout 2019, the annual IRR of first sepsis episode was stable (IRR per year, 0.999; 95% CI, 0.994-1.004), whereas the overall sepsis incidence rate increased (IRR per year increase, 1.013; 95% CI, 1.007-1.019), with a total increase in incidence rates of 15.5%. During the COVID-19 pandemic, the incidence rate was reduced compared to the previously 11-year period, with IRR of 0.877 (95% CI, 0.829-0.927) in 2020 and 0.929 (95% CI, 0.870-0.992) in 2021 for first sepsis cases, and 0.870 (95% CI, 0.810-0.935) in 2020 and 0.908 (95% CI, 0.840-0.980) in 2021 for all sepsis cases. Table 3 shows Poisson regression for trends in first and all sepsis admissions.

### Case Fatality and Temporal Trends

The mean CFR was 13.7% over the fourteen years study period (Figure 3). In-hospital deaths declined during 2009 to 2019 (OR per year, 0.954 [95% CI, 0.950-0.958]), with a total decline of 43.1%. Hospital death increased during the COVID-19 pandemic with an OR 1.061 (95% CI, 1.001-1.124) in 2020 and an OR of 1.164 (95% CI, 1.098-1.233) in 2021 (Table 4).

**Fig. 3.**
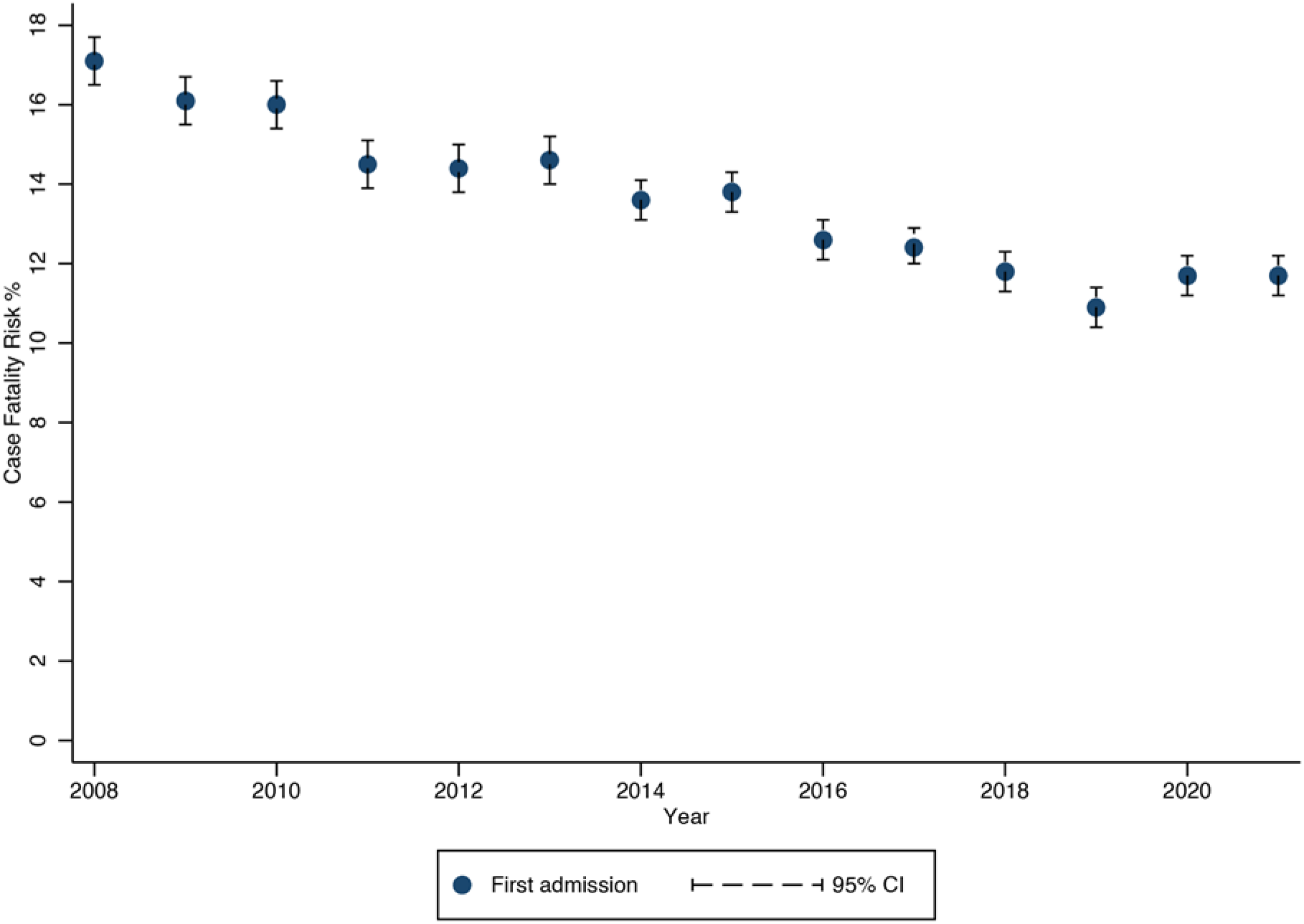
Annual mean case fatality risk in percent among first sepsis admissions (2008-2021) Abbrevation: CI = confidence interval

**Table 4.**
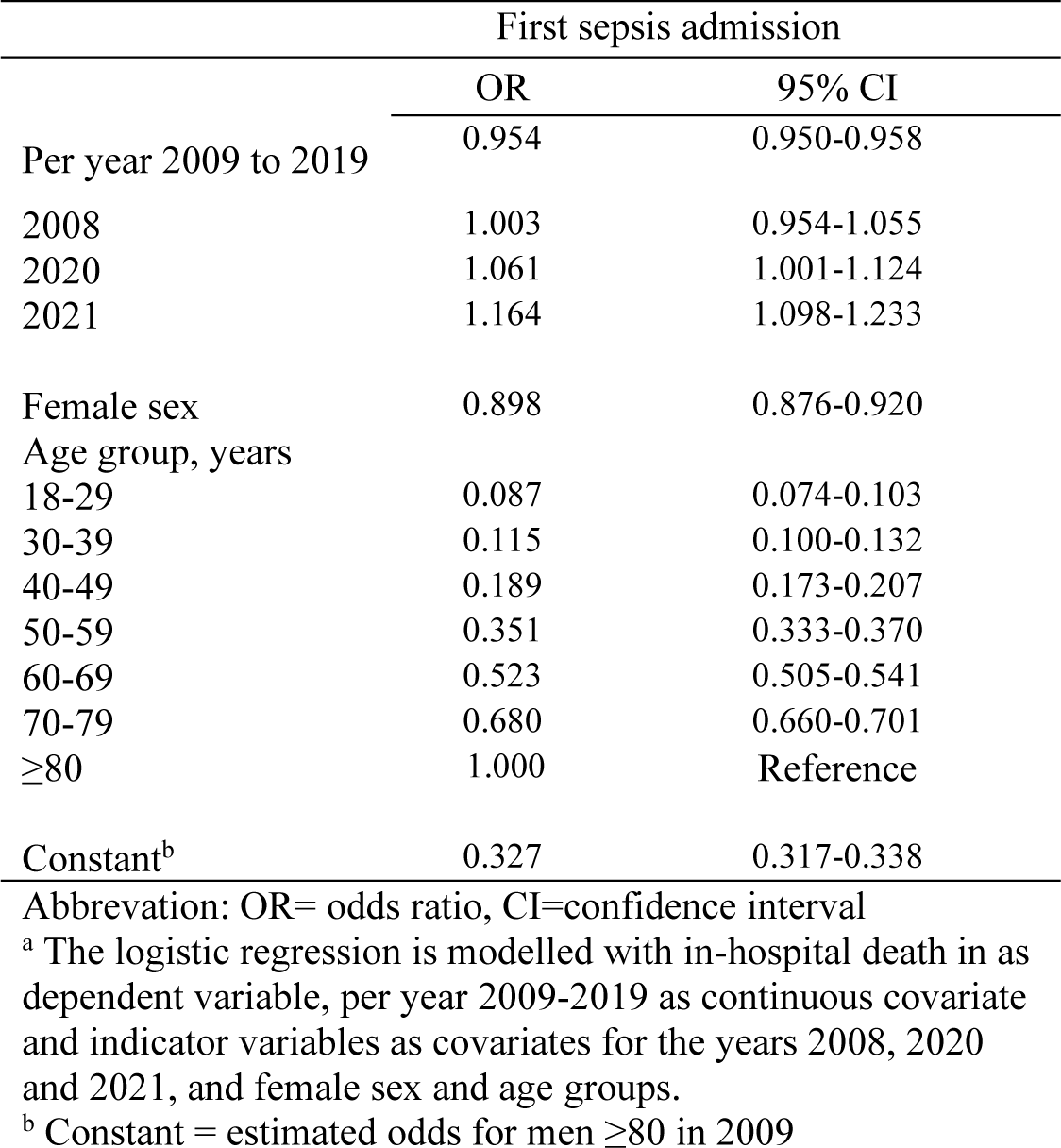
Logistic regression^a^ with in-hospital deaths as dependent variable, 2008-2021.

Quarterly calculations for the years 2020 and 2021 are given in Supplementary Table 3 and Supplementary Figure 3, illustrating that the hospital outcome in COVID-19-related sepsis varied across the pandemic. In contrast, patients with first sepsis admission experienced more stable outcomes over the same period.

## Discussion

In this nationwide longitudinal registry study using all hospital data over fourteen years (2008-2021), we identify a stable trend in the incidence rate of a first sepsis episode but an increasing trend for all sepsis admissions. We also observed a decreasing trend in case fatality. Compared to the period 2009-2019, there was a substantial reduction in sepsis incidence rate during the first year of the COVID-19 pandemic that was somewhat attenuating towards pre-pandemic levels in 2021. Further, we demonstrate an increase in case fatality during the COVID-19 pandemic, most prominent in 2021.

Previously “The Global burden of Disease Study” by Rudd and colleagues (2020) registered an estimated reduction of 37% in the age-standardized incidence rate of sepsis from 1990 to 2017,^2^ and the differences to our study could be due to heterogeneity between regions, the inclusion of low- and middle-income countries with less access to health care, inclusion of persons aged<18 and longer follow-up. Similarities with our study are the use of individual-level data and similar extraction of ICD-10 codes. Several other articles report increasing sepsis incidence rates,^15,17,22,26,27^ i.e., the opposite of what we and Rudd and colleagues found. Martin et al. (2003) found an annual 8.7% increase in sepsis incidence rate using claimed-based data between 1979 and 2000.^26^ Dombrovskiy et al. (2007) found almost doubled hospitalizations of severe sepsis from 1992 to 2003,^17^ and Kumar et al. (2011) calculated an increase in sepsis incidence rate of 200/100 000 inhabitants from 2000 to 2007.^15^ These results are difficult to compare with our analysis regarding first sepsis episodes because they report on all sepsis admissions and fail to stratify on individual entry. However, their results can be compared to our analysis of all sepsis admissions, where we find an increased age-and sex-adjusted incidence rate ratio before the current pandemic. Studies that include all sepsis admissions will naturally increase incidence rates because each person is possibly admitted multiple times, thus increasing the nominator without changing the denominator. Both Rudd and colleagues (2020) and our study go against the myth that the increase in sepsis incidence rates primarily is driven by more liberal practices in sepsis coding over time. It is more likely that previously reported increased incidence rates is caused by the failure to treat each case as an individual entry. Better treatment of medical conditions such as cancer and chronic diseases with increased use of immunosuppressives and invasive procedures ^28,29^ increases the number of patients at risk of developing more than one sepsis episode.^30^ Further, sepsis survivors are prone to recurring sepsis due to new or worsened comorbidities and repeated infections and will thus drive the sepsis nominator.^31^

Previous studies of in-hospital sepsis mortality show in general a decreasing trend. Kaukonen et al. (2014) conducted a retrospective observational study over twelve years of sepsis patients admitted to ICU.^32^ They reported annually decline in mortality throughout the study period with an odds ratio of 0.49 in 2012, with year 2000 as reference. In a European registry-based study of ICU sepsis patients, Yebenes et al. (2017) reported a odds ratio in 2012 with 2008 as reference of 0.77 in a multivariate analysis.^27^ The higher decline than ours can possible be due to inclusion criteria regarding sepsis severity, and that new and updated guidelines, and more attention to the sepsis diagnosis have improved the recognition of the diagnosis, thus assisting clinicians in accurate and timely treatment of infections (i.e., early blood culture sampling and antibiotics), preventing illness severity and therefore reducing mortality.^33-37^

The sepsis incidence rate during the pandemic is previously studied by Bodilsen and colleagues (2021).^14^ They compared hospital admissions for several diagnoses, one year prior to and 11 months after the COVID-19 pandemic and reported a significant reduction in sepsis incidence rate using a few selected sepsis codes and found elevated 30 days mortality.^14^ These previous results are in line with our results. Explanations for the observed lower incidence of sepsis after the pandemic can be the lower incidence of other infections with lockdowns,^14,38^ in addition to vaccination strategies prioritizing the elderly first and canceling elective surgeries.^39^ Other explanations could be a higher threshold for hospitalization during the pandemic in order to avoid an overflow of ill patients to hospitals.

In the above-mentioned Danish study, the 30 days mortality for sepsis under and between the lockdowns was in line with our results.^14^ The increased case fatality in first sepsis admission after the pandemic lockdown can be explained by the fatality of the novel SARS-CoV-2 virus. Further concerns are reluctance to seek health care because of the perceived risk of COVID-19 infection and negligence to report severe symptoms. Probably implications of these explanations are higher in-hospital mortality as those who were admitted with sepsis were more severely ill and thus had a higher baseline mortality risk.

There are several limitations to our study. First, the use of registry-based study design is dependent on ICD-code abstraction and the characteristics of registries.^40^ However, it is mandatory for all Norwegian hospitals to report all activity to NPR and the NPR is a complete and unselected national hospital registry. Identifying sepsis by ICD-10 codes in register-based studies was first used by Angus,^22^ and later modified by Rudd and colleagues to reflect the modern understanding of sepsis pathophysiology.^2^ Different study designs have been investigated to find the most fitted design, with dividing results.^41-44^ The method used by Rudd et al. (2020) has been criticized regarding code selecting strategies that does not fit all countries, and therefore most probable cause an overestimation of sepsis.^45^ The ICD-10 codes are not static, and new specific codes for SIRS and septic shock were implemented in 2010.^46^ We have during the follow-up used the Sepsis-3 definition, albeit the new definition first came in 2016.^1^. However, the trends seem to be consistent across the follow-up period except for 2008 and the pandemic years. Second, the incidence rate of first episodes is probably inflated in 2008, but fitting 2008 as an indicator variable in the regression model will account for this. Third, retrieving organ dysfunction codes to identify implicit sepsis can generate false-positive outcomes since not all organ dysfunctions are caused by a specific infection. On the other hand, false-negative results can occur if the sepsis episode is inadequately documented. Fourth, this study is without an adjustment for illness severity. Our study adjusted for age, and the age differences in sepsis and COVID-19-related sepsis patients can indicate that other demographic characteristics and pathogenesis could affect the association between sepsis, COVID-19-related sepsis, and death. Finally, the influence of the pandemic was calculated from January 2020, although the first COVID-19 patients were first admitted in late February 2020, and thus, the estimated drop in incidence rate related to COVID-19 could be underestimated. It is important to note that the level of SARS-CoV-2 incidence in Norway has been relatively low and therefore, the interpretation of the analysis is primarily relevant to countries with the same burden.

The study also has several strengths, including the large sample size, the use of individual-based data, and a timespan of fourteen years, which makes it possible to detect trends over time. Another strength is that we, in one joint paper, report the burden of first sepsis admissions, all sepsis admissions and case fatality, including age-separated analyses. Since the patients at first admission are likely to be younger, have fewer comorbidities, and thus have less morbidity and mortality risk, stratifying on the first admission will avoid migrating the patient to the next stage, also known as Will Rogers Phenomenon,” or stage migration.^40^. To the best of our knowledge, this is the first study that provides nationwide hospital admissions-based epidemiological characteristics over fourteen years for sepsis and includes data outside the ICU as well as for severe COVID-19-related sepsis.

Our results have implications for health policymakers, clinicians, and researchers. The burden of sepsis is higher than previously described in comparable studies and requires further attention. More sepsis survivors put more pressure on skilled nursing facilities and in-home care. Surveillance and prevention should be assessed and implemented in primary health care. Side-effects of the pandemic, with a pressured healthcare system and a changed threshold for seeking health care, must be evaluated.

## CONCLUSION

This nationwide register-based study over fourteen years reveals that the burden of sepsis still is high. Furthermore, the high incidence rates and decreasing mortality cause an increased number of sepsis survivors, with a growing impact on the healthcare system. Notably, the decreased incidence rates of sepsis hospitalizations together with increased mortality during the pandemics give a concern regarding different efforts that were made to stop the spread of SARS-CoV-2.

## Data Availability

All data produced in the present study are available upon reasonable request to the authors

## Article Information

### Author contributions

*Study concept and design:* Skei, Nilsen, Knoop, Prescott, Damås, Gustad

*Acquisition of data:* Skei, Gustad

*Analysis and interpretation of data:* Skei, Nilsen, Gustad

*Drafting of the manuscript:* Skei, Gustad

*Funding acquisition*: Gustad

*Critical revision of the manuscript for important intellectual content:* All authors

*Statistical analysis:* Skei, Gustad, Lydersen

*Administrative, technical, or material support:* Skei, Brkic, Gustad

*Study supervision:* Nilsen, Damås, Gustad

## Acknowledgments

We would like to thank the Norwegian Patient Registry for data delivery and the clinicians and head of Department of Anesthesia and Intensive Care, Levanger Hospital, HNT HF for support.

