## Supplementary for "Long-term temporal trends in incidence rate and case fatality of sepsis and COVID-19-related sepsis: nationwide registry study"

**Supplementary Table 1** Overview of ICD-10 codes identifying explicit and implicit sepsis

| | |
|---|---|
| Explicit sepsis | A02.1, A20.7, A21.7, A22.7, A24.1, A26.7, A28.2, A32.7, A39.2, A39.4, A40, A41, A42.7, B00.7, B37.7 |
| Implicit Sepsis[a,b] | **Infection**<br>A00-09, A19/28, A30-32, A36/39, A42/ 44, A46, A48/49, A54, A59, A69.0, A69.1, A69.9, A70, A74/75, A77/81, A83/89, A92/99,<br>B00/09, B25/27, B33/34, B37/46, B48/50, B54/55, B57/58, B60, B64, B67, B95/97, B99,<br>G00/08,<br>H05.0, H60.2, H70.0,<br>I00, I33, I38/40.0,<br>J01/06, J09/22, J36, J39.0, J39.1, J85, J86,<br>K35/37, K61, K63.0/63.1, K65, K75.0, K81.0, K83.0,<br>L02/04, L08,<br>M00/01, M86, M72.6,<br>N10, N15.1, N30, N39.0, N41.0, N41.2, N41.3, N45, N70/74, N98.0, N49<br>O03.0, O03.5, O04.5, O08.0, O23, O75.3, O85/86, O88.3, O91, O98,<br>T80.2, T81.4, T82.6/82.7, T83.5/83.6, T84.5/84.7, T85.7, T88.0,<br>U04, U07.1, U07.2<br><br>AND<br>**Acute organ dysfunction**<br>D65, D69.5, E87.2, G93.4, I46, I95.9, J80, J95.2, J96, K72.0, K72.9, N00, N17, N99.0, R02, R09.0, R09.2, R40.0/40.2, R41, R55, R57, R57.2, R65.1[c] |

Abbreviation: ICD= International Classification of Diseases

[a] Implicit sepsis was defined if one code of infection was present with at least one acute organ dysfunction within same hospital entry. Total sepsis estimates are calculated from both explicit and implicit cases.

[b] Explicit codes are excluded from infection codes

[c] If R65.1 was included in the count of acute organ dysfunctions if present in combination with another specific acute organ dysfunction code

**Supplementary Table 2** ICD 10 codes identifying comorbidities and infection sites.

| **Comorbidities** | **ICD-10** code |
|---|---|
| Chronic heart- and vascular disease | G45, H34, I00/31, I34/37, I42/45, I47/95.8, I97/99 |
| Cancer | C00/97, D32/33, D35.2/35.4, D42, D43, D44.3/44.5, D45/47 |
| Chronic lung disease | J41/47, J84, J98 |
| Chronic renal disease | N18.3/18.5 |
| Diabetes | E10/11 |
| Dementia | F00/03, G30, G31.0, G31.2, G31.8 |
| Chronic immune disease | D80/84, Z94.0/94.4, Z94.8 |
| Chronic liver disease | K70.4, K72 |
| **Infection sites*** | |
| Respiratory | J09/18, J20/22, J85/86, U04, U07.1, U07.2 |
| Genitourinary | N10, N15.1, N30, N39.0, N41.0, N41.2/41.3, N45, N49, N70, N71/74, N98.0 |
| Intra-abdominal | A00/09 |
| Extra-abdominal | K35/37, K57, K61/61.1 K61.3, K63.0/63.1, K65, K75.0, K81.0, K83.0 |
| Endocarditis/myocarditis | I32/33, I38/41 |
| Skin/ Soft tissue | A46, B08/09, L02/04, L08, M72.6 |
| Infection after procedure | T80.2, T81.4, T82.6/82.7, T83.5/83.6, T84.5/84.7, T85.7, T88 |
| Other* | A19/28, A30/32, A36/39, A42/44, A48/49, A54, A59, A69.0, A69.1, A69.9, A70, A74/75, A77/80, A81, A83/89, A92/B06, B25/27, B33/34, B37/46, B48/50, B54/55, B57/58, B60, B64, B67, B95/97, B99, G00/08, H05.0, H60.2, H70.0, J01/06, J36, J39.0/39.1, M00/01, M86, O03.0, O03.5, O04.5, O08.0, O23, O75.3, O85/86, O88.3, O91, O98 |
| **Acute organ dysfunction** | |
| Respiratory | J80, J95.2, J96, R09.0, R09.2 |
| Circulatory | I46, I95.9, R57, R57.2 |
| Renal | N00, N17, N99.0 |
| Hepatic | K72.0, K72.9 |
| Coagulation | D65, D69.5 |
| Other acute organ dysfunctions | G93.4, R40.0/40.2, R41, R55, E87.2, R02, R65.1 |

*Explicit codes are excluded from infection codes and other infection sites

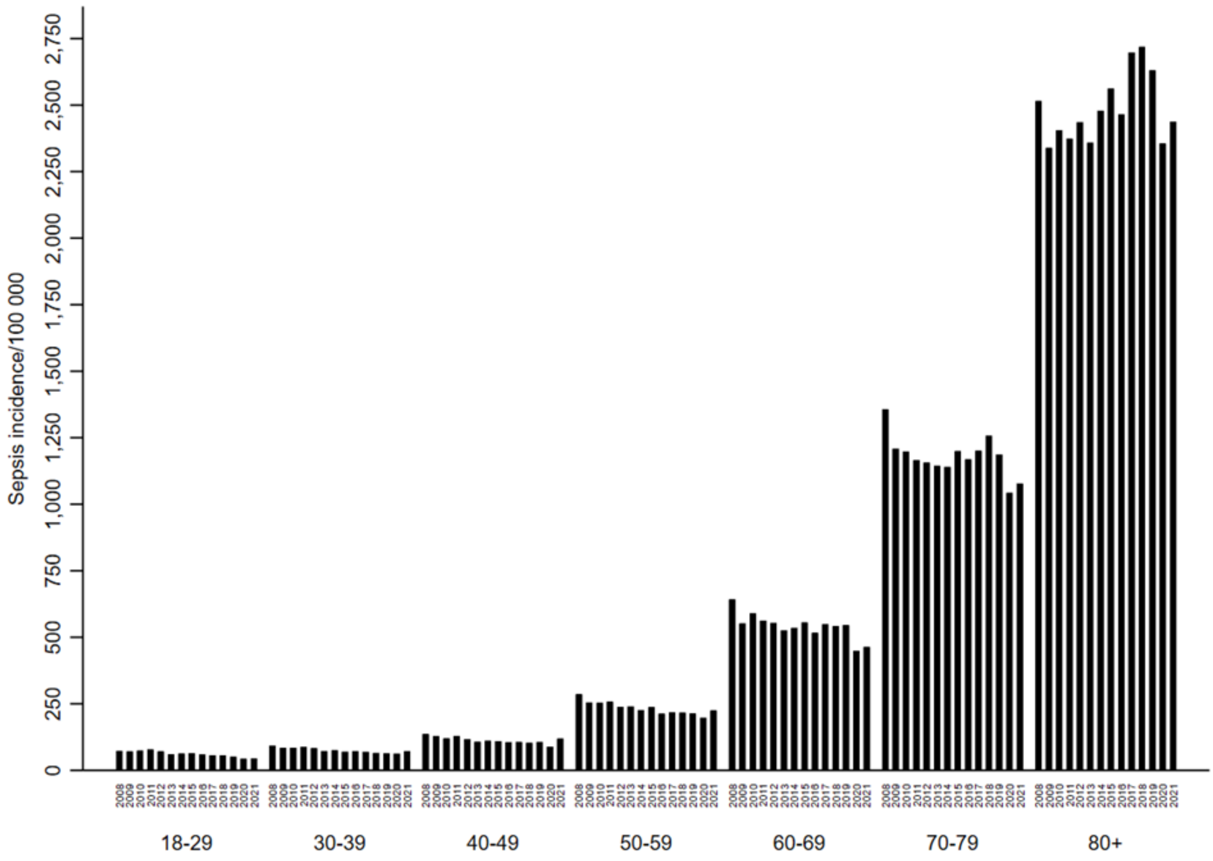


*Supplementary Fig 1 Annual sepsis incidence rates for first admission by ten-years age groups*

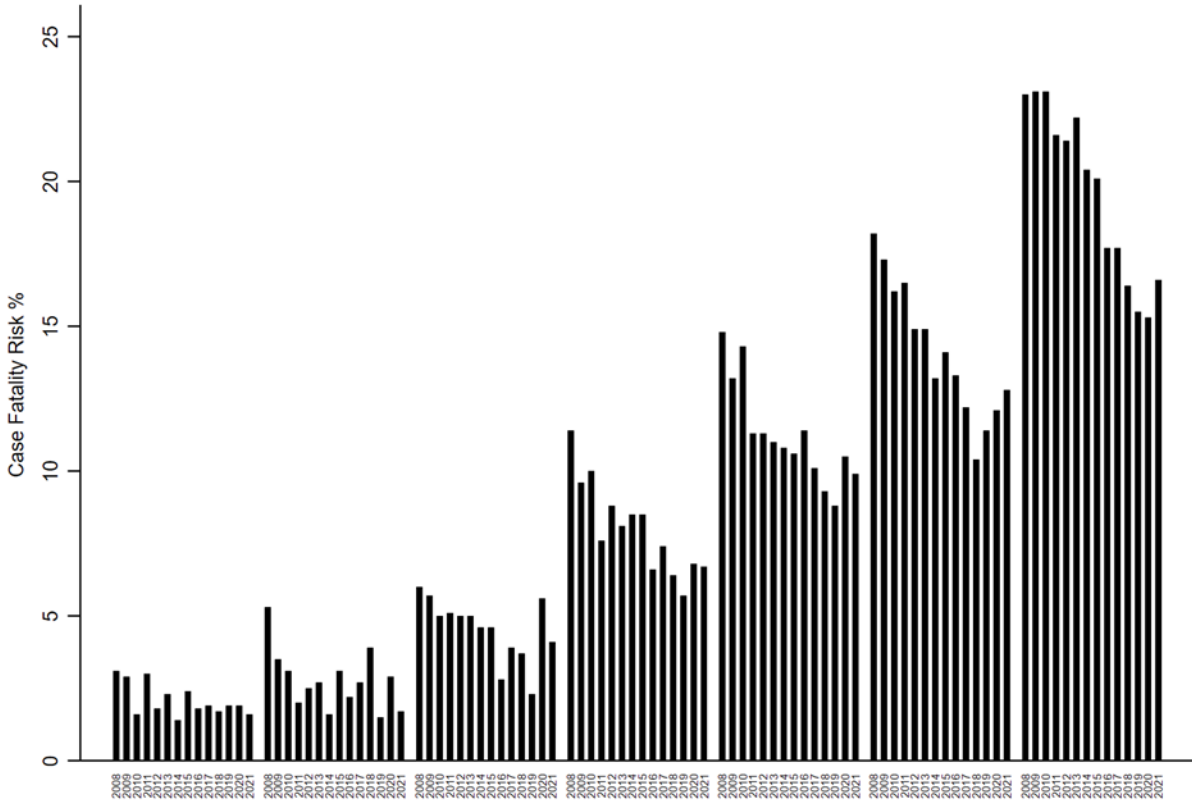


*Suppl Fig 2 Annual case fatality risk in % for first sepsis admissions by ten years age-groups*

**Supplementary Table 3** First admissions, deaths, and CFR for sepsis and COVID-19-related sepsis patients in 2020 and 2021.

| | 2020 | | | | | | 2021 | | | | | |
|---|---|---|---|---|---|---|---|---|---|---|---|---|
| | Sepsis | | | COVID-19-related sepsis | | | Sepsis | | | COVID-19-related sepsis | | |
| | N | Deaths | CFR % | N | Deaths | CFR % | N | Deaths | CFR % | N | Deaths | CFR % |
| Q1 | 4310 | 505 | 11.7 | 266 | 42 | 15.8 | 3335 | 415 | 12.4 | 655 | 58 | 8.9 |
| Q2 | 3140 | 371 | 11.8 | 166 | 23 | 13.9 | 3336 | 401 | 12.0 | 389 | 25 | 6.4 |
| Q3 | 3501 | 384 | 11.0 | 54 | 5 | 9.3 | 3734 | 446 | 11.9 | 225 | 32 | 14.2 |
| Q4 | 3720 | 438 | 11.8 | 290 | 39 | 13.4 | 4233 | 505 | 11.9 | 800 | 128 | 16.0 |

Abbreviations: N = Number of cases, CFR= Case Fatality Risk calculated as in-hospital death divided by first sepsis admission in the quarter (Q). Q1 (January, February, March), Q2 (April, May, June), Q3 July, August; September, Q4 (October, November, December).

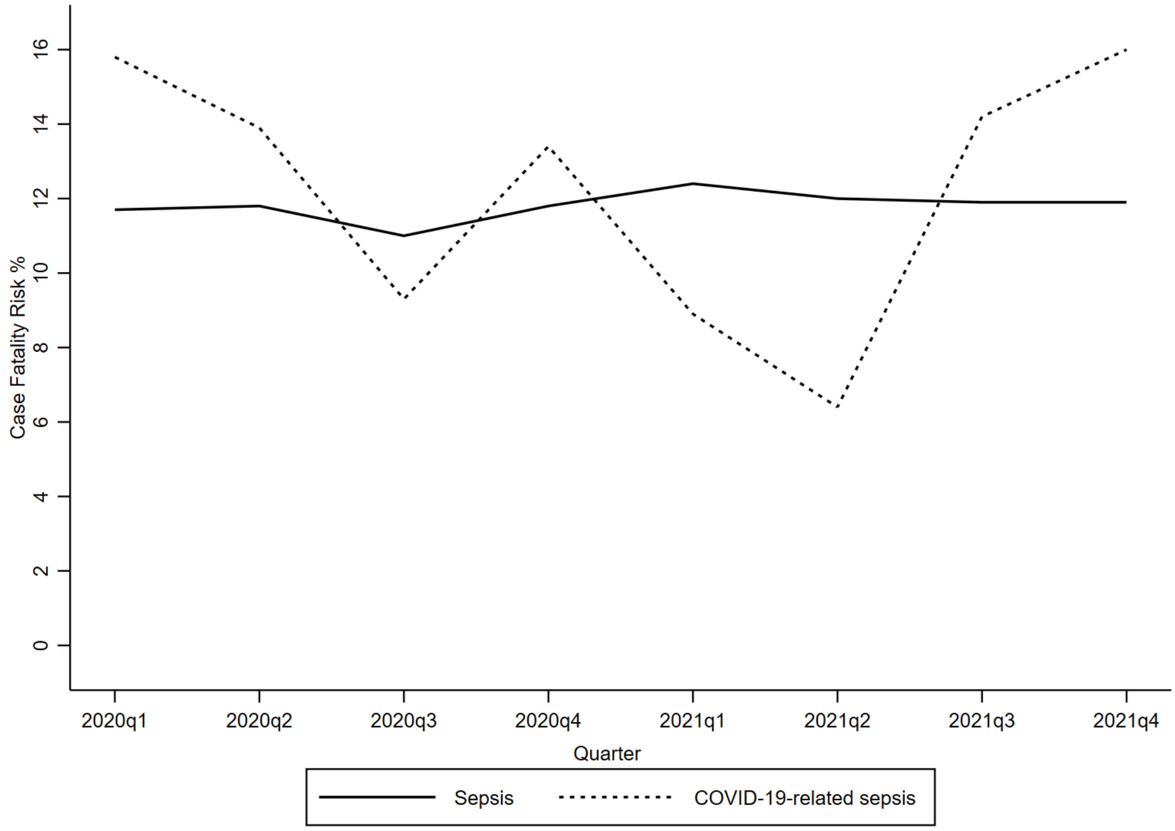


*Note*: Calculated as **Q1** (January 2020, February 2020, March 2020), **Q2** (April 2020, May 2020, June 2020), **Q3** (July 2020, August 2020, September 2020), **Q4** (October 2020, November 2020, December 2020), **Q1** (January 2021, February 2021, March 2021), **Q2** (April 2021, May 2021, June 2021), **Q3** (July 2021, August 2021, September 2021), **Q4** (October 2021, November 2021, December 2021).

*Supplementary Fig 3 Quarterly mean case fatality risk in sepsis and COVID-19-related sepsis for first admission (2020 and 2021)*
